# Bioinformatics and machine learning-based identification of cell cycle-related genes and molecular subtypes in endometrial cancer

**DOI:** 10.1101/2024.11.27.24318050

**Authors:** Jingying Pan, Shuhan Huang, Bidong Fu, Ruiyu Zhang, Minqin Zhou, Zichuan Yu, Hong Zeng, Xitong Geng, Yanting Zhu, Hao Zheng, Hao Wan, Xiaoyu Qu, Shengwei Tang, Yanying Zhong

**Affiliations:** Department of Obstetrics and Gynecology, Second Affiliated Hospital of Nanchang University, Nanchang, China; First College of Clinical Medicine, Nanchang university, China; Second College of Clinical Medicine, Nanchang university, China; HuanKui Academy, Nanchang University, China; School of Ophthalmology&optometry,Nanchang University, China

## Abstract

Endometrial cancer is a common malignant tumor in women, with rising incidence rates and an unoptimistic prognosis. DSN1 is a kinetochore protein-coding gene that affects centromere assembly and progression in cell cycles, which is associated with adverse predictions for many cancers. However, the role of DSN1 in UCEC has not yet been reported. We identified the UCEC-related gene module and obtained the differential genes. Then we constructed a diagnostic model and identified the subtype of the molecule and its association with predictions. Subsequently, we identified DSN1 as the core gene and predicted its predictive value. Furthermore, using bioinformatics methods, we found DSN1 was associated with certain clinical characteristics and experimentally validated the expression in cancer tissues of DSN1. Pathway enrichment analysis identified DSN1 as a cell cycle-associated protein, which was validated by WB. The protein interaction network also revealed DSN1 was significantly associated with NDC80. Then we explored the correlation of DSN1 and immune cells and immune cell infiltration and found that DSN1 may affect Th2 enrichment by affecting CCL7 and CCL8. Drug susceptibility analysis showed DSN1 was sensitive to cisplatin and resistant to sunitinib. In conclusion, DSN1 was a novel biomarker that contributes to prognosis and treatment.

## Introduction

Endometrial cancer (UCEC) is the fourth most familiar cancer in women (1). The morbidity of UCEC has increased by 132% in the last three decades, and the mortality rate has also increased (2, 3). Most patients with early-stage endometrial cancer are relatively manageable by the hysterectomy and adjuvant radiotherapy (4). However, the current prognosis for terminal UCEC patients remains poor, with a five-year survival rate of only 15% for stage IV patients (5). Although chemotherapy is the standard treatment for patients with terminal or relapsing endometrial cancer, the outcome is unsatisfactory (6). So there is still a need to find ways to assist in pre-diagnosis to ameliorate the prognosis of UCEC. Biomarkers play a significant part in the identification, early diagnosis, and disease prevention and monitoring during treatment. Therefore, it is significant to find biomarkers that can enhance the prognosis of endometrial cancer and assist in early diagnosis.

DSN1(DSN1 component of MIS12 kinetochore complex) is a widely distributed protein in the centromere, and its coding gene is located on chromosome 20q11.23. DSN1 plays a vital part in many biological processes, for example, mitosis and cell cycle. Some studies have indicated that chromosomal instability is connected with the high expression of DSN1. And chromosomal instability can lead to chromosomal structural and quantitative abnormalities, which is a prominent feature of human cancers. DSN1 has been reported to have profound significance in the progression of various tumors, including breast carcinoma (7), colorectal carcinoma (8), hepatocellular carcinoma (9), and sarcoma (10). For example, in colorectal and hepatocellular carcinomas, DSN1 affects cell cycle progression and is closely related to its clinicopathological features, and in vitro, DSN1 significantly promotes osteosarcoma cell proliferation (11). However, the function of DSN1 in UCEC has not been reported.

With the bioinformatics and experimental method, we investigated the potential function and mechanism of DSN1 in UCEC. By the LASSO regression and consensus cluster analysis, we confirmed the core genes and determined the molecular subtypes of UCEC and their relationship with prognosis. Then, survival and random forest analysis were utilized to identify the key gene, DSN1. The expression of DSN1 in UCEC and its correlation with prognosis were analyzed by R software, qRT-PCR, and Western Blot experiments. Subsequently, Linkedomics and GSEA were utilized to investigate the pathway enrichment and latent biological functions of DSN1 and verified by WB. Using STRING, we constructed a DSN1 protein interaction network. In addition, the relationship between DSN1 and tumor-infiltrating immune cells, chemokines, and drug interactions in UCEC were also investigated. Our study suggested that DSN1 is a novel biomarker of UCEC that contributes to prognosis and treatment.

## Materials and methods

### Clinical samples

The specimens of all 50 patients from the Second Affiliated Hospital of Nanchang University were pathologically identified with UCEC between December 2023 and September 2024. While the tissues were still fresh, total proteins were removed. The study was approved by the ethics committee of Nanchang University’s Second Affiliated Hospital (No. Review [2023] No. (165)). This is a retrospective study. As it analyzed existing medical records and archived samples, obtaining informed consent was infeasible. The ethics committee of Nanchang University’s Second Affiliated Hospital has approved the waiver of informed consent for this clinical research. And after the data is collected, information can be obtained that can identify individual participants.

### Data Collection and Proceed

Sequencing and clinicopathological data of UCEC patients were derived from the TCGA database (https://cancergenome.nih.gov), including 59 normal samples and 554 tumor samples. The mRNA expression profiles (Number: GSE17025, GSE39099, and GSE106191) were derived from the GEO database (https://www.ncbi.nlm.nih.gov/geo/).

### Determination of differentially expressed genes (DEGs)

The gene sets were normalized, and difference analysis was carried out using the “limma” package to get DEGs between each comparison group and the control group. We used a P-value less than 0.05 and |log2 fold change (FC) |> 1 to determine DEGs.

### Cell culture

The commercially accessible UCEC cell line HEC-1-B was utilized to matrix cells, while the detailed method can be referred to in this article (12).

### Transfection with shRNA

Specific sequences were designed to manipulate the expression of DSN1. ShRNAs were cloned into the lentiviral pLKO.1 vector, which was purchased commercially from GenePharma Biothch in Shanghai, China.

### RNA extraction and qRT-PCR

The TRIzol reagent (Invitrogen, USA) per the manufacturer’s instructions was applied to isolate total RNA from the cells. Invitrogen’s M-MLV reverse transcription kit was used to do reverse transcription on total RNA. The Agilent Technologies AriaMx Real-Time PCR System (Agilent, USA) and SYBR Green PCR Master Mix (Roche, Switzerland) were utilized for the qRT-PCR analysis. The relative expression was computed using the 2 (delta delta threshold cycle) (2Ct) technique with tubulin as a normalized internal control.

### Western blotting analysis and antibodies

Western blotting analysis was carried out. Primary antibodies included anti-DSN1 (1:1000, Cell Signaling Technology 4724), anti-NDC80 (1:1000, Santa Cruz, sc-515550), anti-Cyclin D1 monoclonal antibody (1:1000, Abcam, ab16663), anti-Tubulin monoclonal antibody (1:1000, Proteintech, 11224-1-AP).

### WGCNA (Weighted correlation network analysis)

Using WGCNA, we uncovered interesting gene modules that are closely connected to other genes. First, transpose the concatenated matrix file, then screen out the genes with expression variance in the first quartile before constructing the correlation matrix. The weighted adjacency matrix is then transformed into a topological overlap matrix (TOM) to assess network connectivity, and the hierarchical clustering approach is utilized to create the TOM clustering tree. Genes were sorted into modules based on their expression patterns using a weighted correlation coefficient. The relationship between modules and clinical characteristics was further investigated to identify modules linked with UCEC for investigation.

### Gene function enrichment analysis

GSEA was implemented by applying R package ClusterProfiler 4.1.0, and genomic enrichment analysis of differentially expressed genes of TCGA and GEO was performed. Prominent functional and pathway differences between high and low DSN1 expression groups were also analyzed by GSEA. Based on gene expression profiles and phenotypic grouping, we kept the minimum gene set at 5 and the largest gene set at 5,000. p values less than 0.05 and FDR less than 0.1 were deemed statistically significant. Then, R packages “clusterProfiler” and “org.hh.egg.db” were applied to gene ontology (GO) and Kyoto Encyclopedia of Genes and Genomes (KEGG) analysis to further analyze the associated pathways of 164 genes and 20 genes in UCEC. An adjusted p-value of less than 0.05 was considered significantly enriched.

### Machine learning identification of candidate genes associated with UCEC

Using the least absolute shrinkage and selection operator (LASSO), the final core genes were screened. A diagnostic model was applied to determine a risk score for each patient. The random forest algorithm is an algorithm based on bagging. Bagging is an integration method that randomly divides a data set into multiple subsets to build a basic decision tree. We used a random forest algorithm to score these genes.

### Identification of molecular-related subtypes by consensus clustering

The samples are categorized by determining the consensus matrix through consensus clustering, and the K-value for the number of clusters is set between 2 and 9. The optimal K value was determined when the cumulative distribution function index reached an approximate maximum. Correlated subtypes were established based on consensus clustering of 20 gene expression levels. These results were used to define Cluster 1, Cluster 2, and Cluster 3. Heat maps were plotted using the ggplot2 software package to visualize the expression of these 20 genes in the different clusters.

### LinkedOmics Database Analysis

The LinkedOmics database (http://www.linkedomics.org/login.php) was used to evaluate 32 TCGA cancer-related datasets. Its “LinkFinder” module has been used to investigate differential expression genes associated with DSN1 in the TCGA UCEC. If the P-value and false discovery rate are both less than 0.05, the gene will be considered a significant related gene.

### Protein-protein interaction network construction

The STRING database (https://string-db.org) allows us to explore the genomic relationships between genes encoding proteins and thus infer the biological activity of proteins. Our study utilized the STRING database to explore the top 500 co-expressed DSN1 genes and selected medium-confidence genes with a confidence level of 0.9, which were then displayed by Cytoscape software to generate an overall PPI network.

### TIMER Database Analysis

The Tumor Immune Estimation Resource (TIMER) database could provide immune infiltration information for different cancer types systematically. We utilized “Gene”, “Diff Exp”, and “Correlation” modules to acquire immune-related information and gene expression level information.

### TISIDB analysis

The TISIDB database (http://cis.hku.hk/TISIDB/) could be used to investigate the association between individual genes and immunity in the setting of cancer. Using the “Chemokine” module, we investigated the link between DSN1 expression and cytokines in UCEC.

### Comparative toxicogenomics database

The CTD (http://ctdbase.org/) integrates the interactions of specific genes with relevant compounds and contains literature coverage of a number of compounds. From this, we got several drugs that regulate DSN1 expression.

### Statistical Analysis

In this study, all statistical studies were accomplished by R software (version 3.6.3/4.0.3/4.1.2).

### Survival and clinicopathological features analysis

The R packages involved are “pROC”, “timeROC” and “ggplot2”. By using logistic regression, we investigated the association between DSN1 and clinicopathological features. In addition, DSN1 and the prognostic model use the “Survival” software package for survival regression fitting. And survival analysis for different subgroups. Then, to compare various survival factors, our team developed time-dependent receiver operating characteristic curves (ROC) and used the “rms” package to build a nomogram model and visualize it.

### Correlation analysis

Using the R tools (limma, ggplot2, and heatmap packages), we conducted gene expression correlation analysis. The correlation of DSN1 with DSN1-related genes. The correlation of CCL7 and CCL8 with Th2 cells.

### Calculation of immune infiltration

We used R-packet GSVA and 24 immune cell markers to calculate the immune infiltration corresponding to DSN1.

## Result

### Modular cluster analysis of endometrial cancer-related genes using weighted gene co-expression network analysis (WGCNA)

Aimed to explore critical modules and genes linked with endometrial cancer (UCEC), we used the mRNA expression matrix of TCGA-UCEC, GSE17025, GSE39099, and GSE106191. First, we identified 3922 and 9037 differential genes for GEO (P-value <0.05) and TCGA (P-value<0.05, FDR<0.05, and |logFC|>1)(Figure 1A-B), which were then used for functional enrichment analysis and WGCNA. The GSEA analysis revealed significant links between UCEC-related differential genes and cell cycle regulation and immunological pathways (Figure 1C, D). We then separately constructed the WGCNA model and identified the gene co-expression modules (Figure 1E, F). We found that the yellow modules containing 371 genes were most relevant to the UCEC for the TCGA-UCEC database, while it contained 429 genes for the GEO. Furthermore, we conducted a cross-analysis of the genes and obtained a set of 164 genes (Figure 1G), which was analyzed by GO and KEGG, showing that these genes were closely related to DNA replication, cell cycle, and p53 signaling pathways (Figure 1H, I). Furthermore, random forest algorithms showed higher scores for the 15 genes with higher predictive values in the gene concentration (Figure 1J). Overall, through a comprehensive analysis of multiple datasets, we identified modules and genes that were closely related to UCEC, and they were dramatically gathered in cell cycle-related pathways.

**Figure 1.**
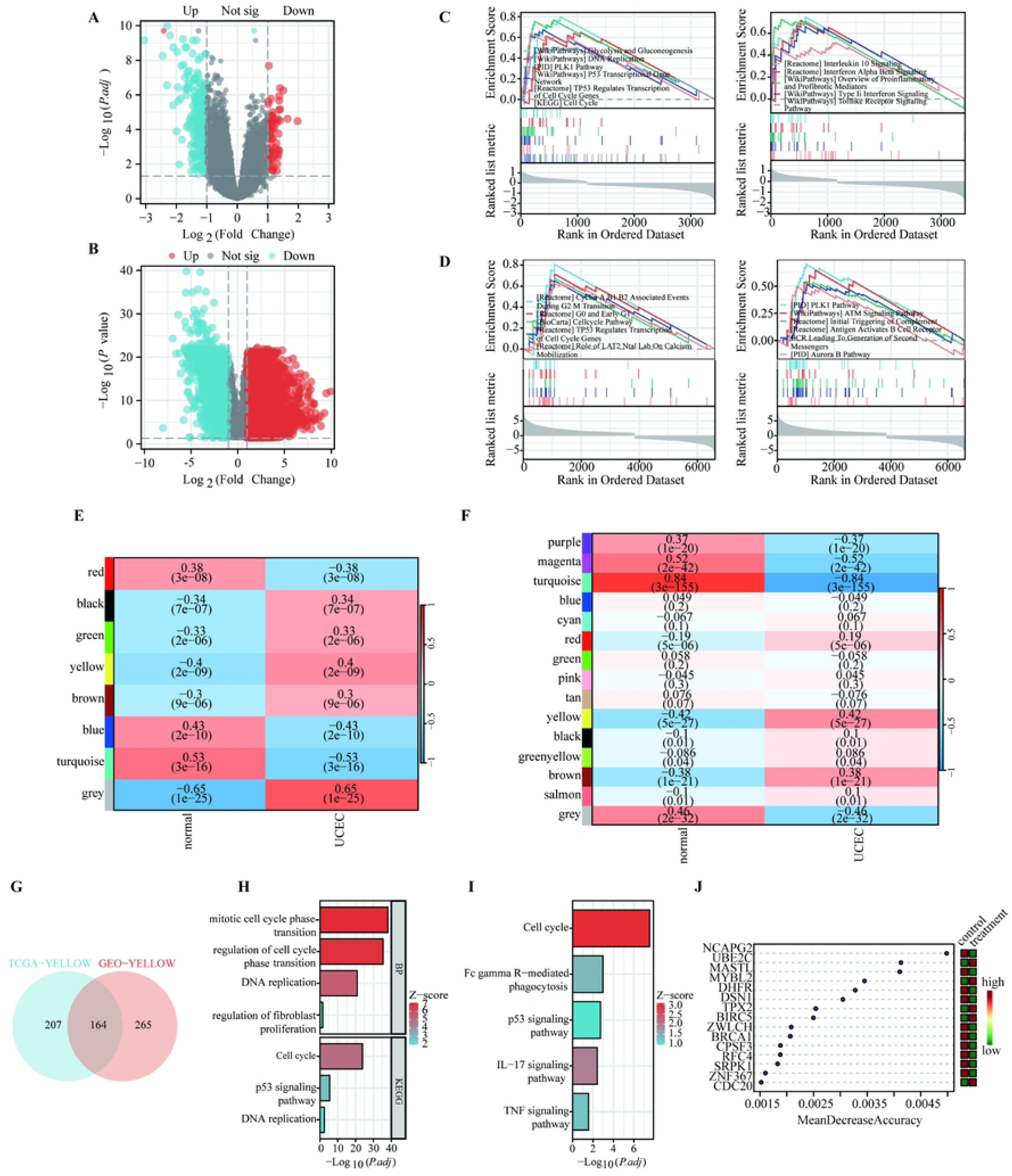
Identification of key gene modules in WGCNA. (A-B) The volcano map of the DEGs screening from the TCGA-UCEC and GSE17025, GSE39099, and GSE106191 data sets. (C-D) GSEA analysis of DEGS. (E) WGCNA analysis identified the module related to UCEC in the TCGA database as the yellow module (r = 0.4). (F) WGCNA analysis confirmed the module most associated with UCEC in the GEO dataset as the yellow module (r = 0.42). (G) The Venn figure depicted the TCGA-UCEC, GEO-UCEC intersection of related genes. (H, I) The GO and KEGG pathway analysis of the gene set containing 164 genes. (J) The random forest analysis.

### LASSO regression analysis further built a diagnostic model containing 20 genes

To build the predictive model and improve the accuracy of the prediction, we employed an LASSO regression analysis of the 164 genes. Then, we refined 20 core genes (Figure 2A, B). Simultaneously, they had higher levels of expression in cancer tissue than normal tissue (Figure 2C). Based on these key genes, we constructed a diagnostic model whose ROC curve analysis AUC values reached 1, showing high diagnostic accuracy (Figure 2D). GO and KEGG pathway analyses were used to understand the biological functions of genes, showing that the genes were highly concentrated in the pathways related to cell cycle regulation (Figure 2E).

**Figure 2.**
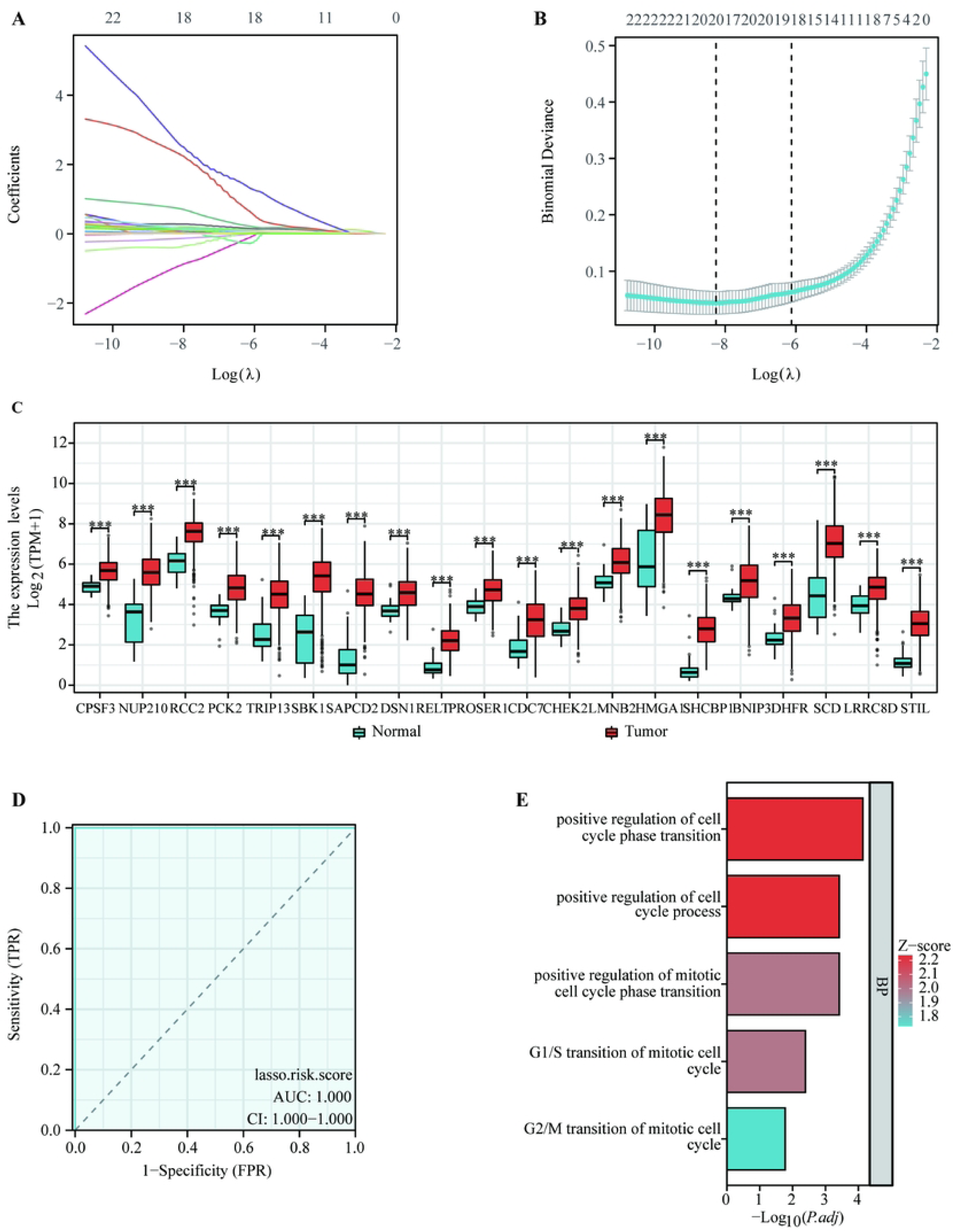
A LASSO regression diagnosis model was constructed. (A-B) LASSO regression analysis was used to determine the most critical model genes, and a new gene set containing 20 genes was obtained. (C) Expression profiles of 20 genes. (D) Diagnostic ROC curves of key gene sets in UCEC. (E) GO and KEGG pathway enrichment analysis of gene sets.

### Identification of molecular subtypes of endometrial cancer-related genes

To study the biological differences between the different subtypes of UCEC, the ConsensusClusterPlus package was used to conduct a consistent clustering analysis of the expression profiles of 20 model genes. CDF curve results showed that when k = 3, the classification was dependable and steady (Figure 3A-C). Meanwhile, the specific expression of the 20 genes in the three clusters was presented in the hot chart (Figure 3D). Immediately, we compared the survival conditions of these three clusters (C1, C2, and C3), suggesting that the C3 cluster had the worst prognostic effect and that there were no significant differences between C2 and C1 (Figure 3E). Overall, there may be some differences in prognosis between the different subtypes of these UCEC-related genes.

**Figure 3.**
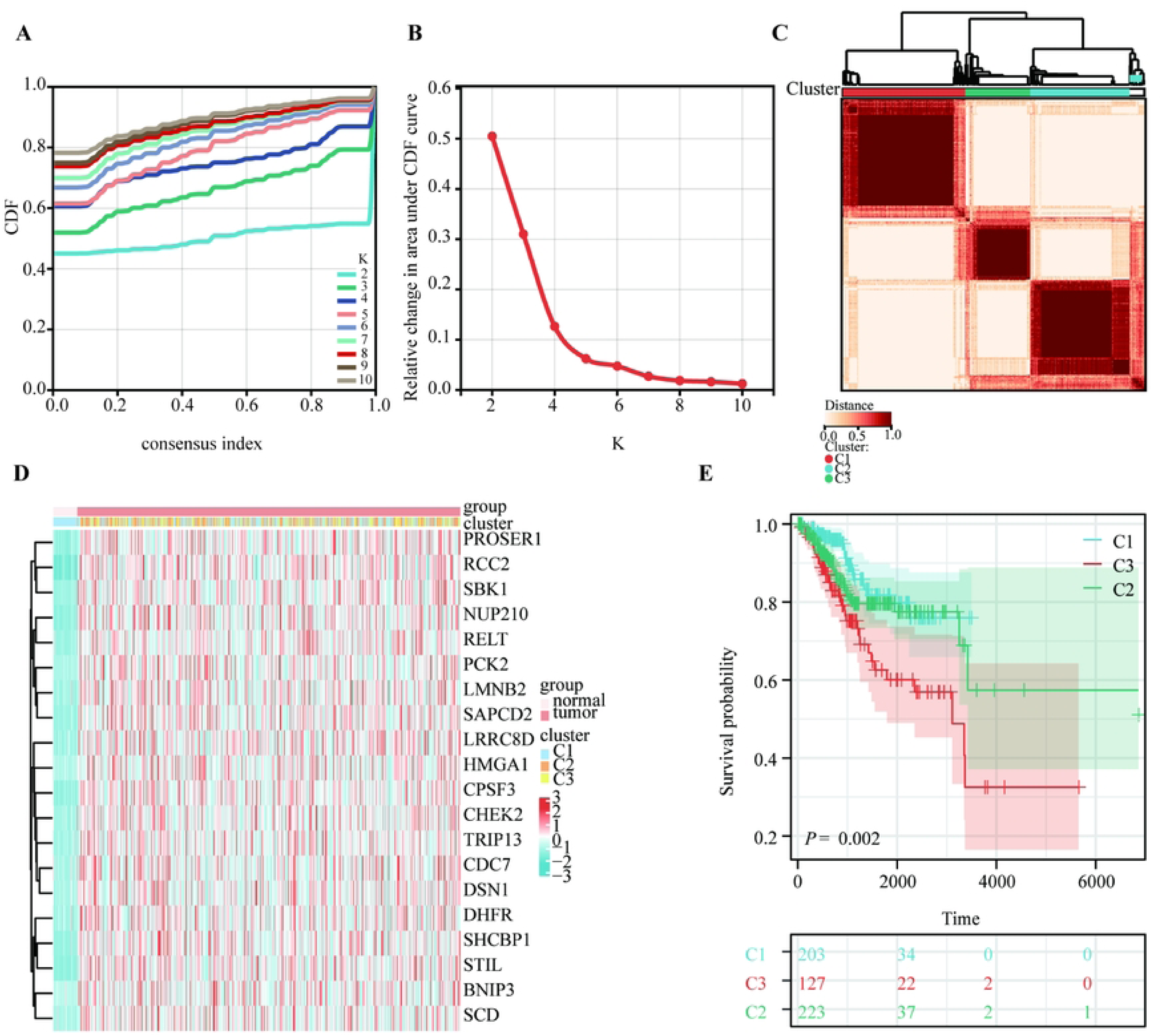
Identification of the molecular subtypes of genes associated with endometrial cancer. (A-C) Consensus clustering determined three UCEC clusters with different enrichment scores. (D) A heat map depicting the expressions of 20 genes in the three clusters. (E) The UCEC OS curve of patients between different subtypes.

### DSN1 had the highest research value based on 20 gene survival analyses

To explore the predictive and diagnostic value of the genes, we used R software to map the survival curves. It indicated that the eight genes, such as DSN1, had a significant and strong correlation between their high levels of expression and the patient’s poor overall survival (OS), suggesting that they may be potential biological markers for disease diagnosis and prognosis assessment (Figure 4A). In contrast, the results of the remaining 12 genes showed no statistically significant differences (as shown in Supplementary Figure 1A). Furthermore, according to the random forest chart score (Figure 1J), the DSN1 gene had the highest score, and its specific role in UCEC had not yet been explored, which greatly aroused our interest. So we further explored its potential biological function and its role in the cancer process. The diagnostic ROC curve of DSN1 showed a high diagnostic value of AUC of 0.966 (Figure 4B). We further constructed a nomogram containing the parameters of the age and different expressions for DSN1. And the calibration chart validates the calculated scores that can accurately predict the 1-, 3-, and 5-year survival chances (Figure 4C, D).

**Figure 4.**
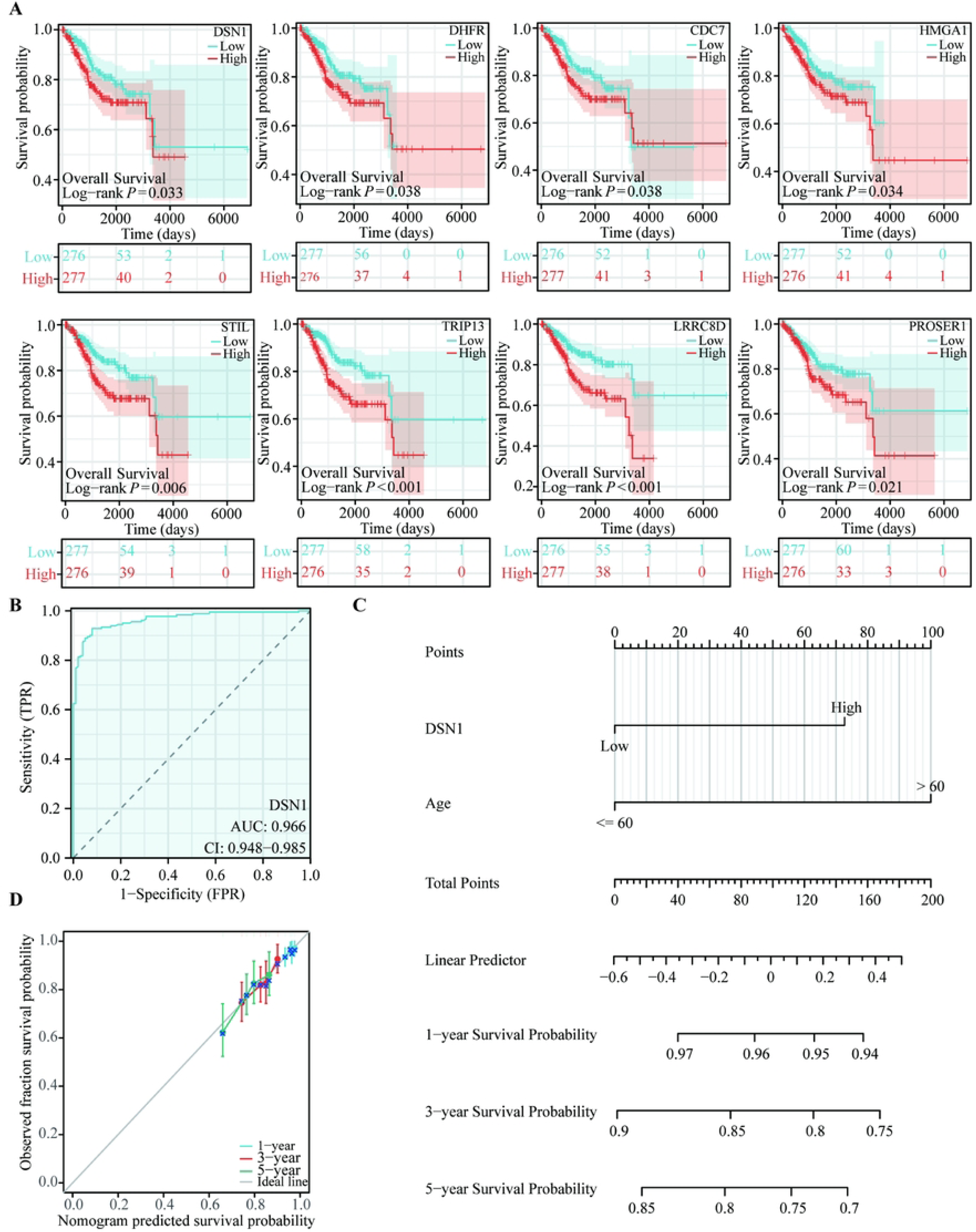
Survival analysis of model genes and identification of core genes. (A) OS curves of model genes with statistically significant survival. (B) Diagnostic ROC curve of DSN1 expression in endometrial cancer. (C) Nomogram to predict 1-, 3-, or 5-year OS rates in patients with endometrial cancer. (D) Calibration curve of the nomogram.

### DSN1 was associated with multiple clinical pathology characteristics in UCEC

To explore the link between DSN1 and the clinical characteristics in UCEC, we conducted gene expression and clinical pathological characteristics analysis. It showed the expression of DSN1 in tumor tissue was significantly higher (P < 0.001) (Figure 5A, B). Next, we detected the links between gene expression and clinical pathological characteristics. In terms of OS, significantly increased DSN1 was observed in UCEC-dead patients (P < 0.05) (Figure 5C). Meanwhile, vitally increased DSN1 was observed in the group over 60 (Figure 5D). As the tumor developed, the expression of DSN1 also increased significantly with the increase in histologic grade (Figure 5E). In the clinical stage, DSN1 expression in Stage I was prominently lower than that in Stage III (Figure 5F). Subsequently, we mapped the total survival curve under different clinical pathological characteristics, assessing their prognosis and diagnostic value. It demonstrated that the overall survival rate was lower with high expression of DSN1 for a worse prognosis, no matter if the group was for different ages (Figure 5G, H). The OS for patients with high expression of DSN1 was low in prognosis at G1 and G2, while G3 was without significant differences (Figure 5I, J). The OS for patients with high expression of DSN1 was low in prognosis at Stage I and Stage II, while at Stage III and Stage IV, there were no significant differences (Figure 5K, L). The qRT-PCR was performed on the tumor tissues and corresponding non-tumor tissues of 50 UCEC patients, and it showed that the mRNA expression levels of 33 of these samples were significantly upregulated (Figure. 5M). According to the results of WB, the experiment DSN1 protein levels were significantly increased in cancer tissue (Figure 5N, O). In conclusion, the expression of DSN1 was significantly increased in cancer tissues, and its differential expression may have different effects under different clinicopathological characteristics.

**Figure 5.**
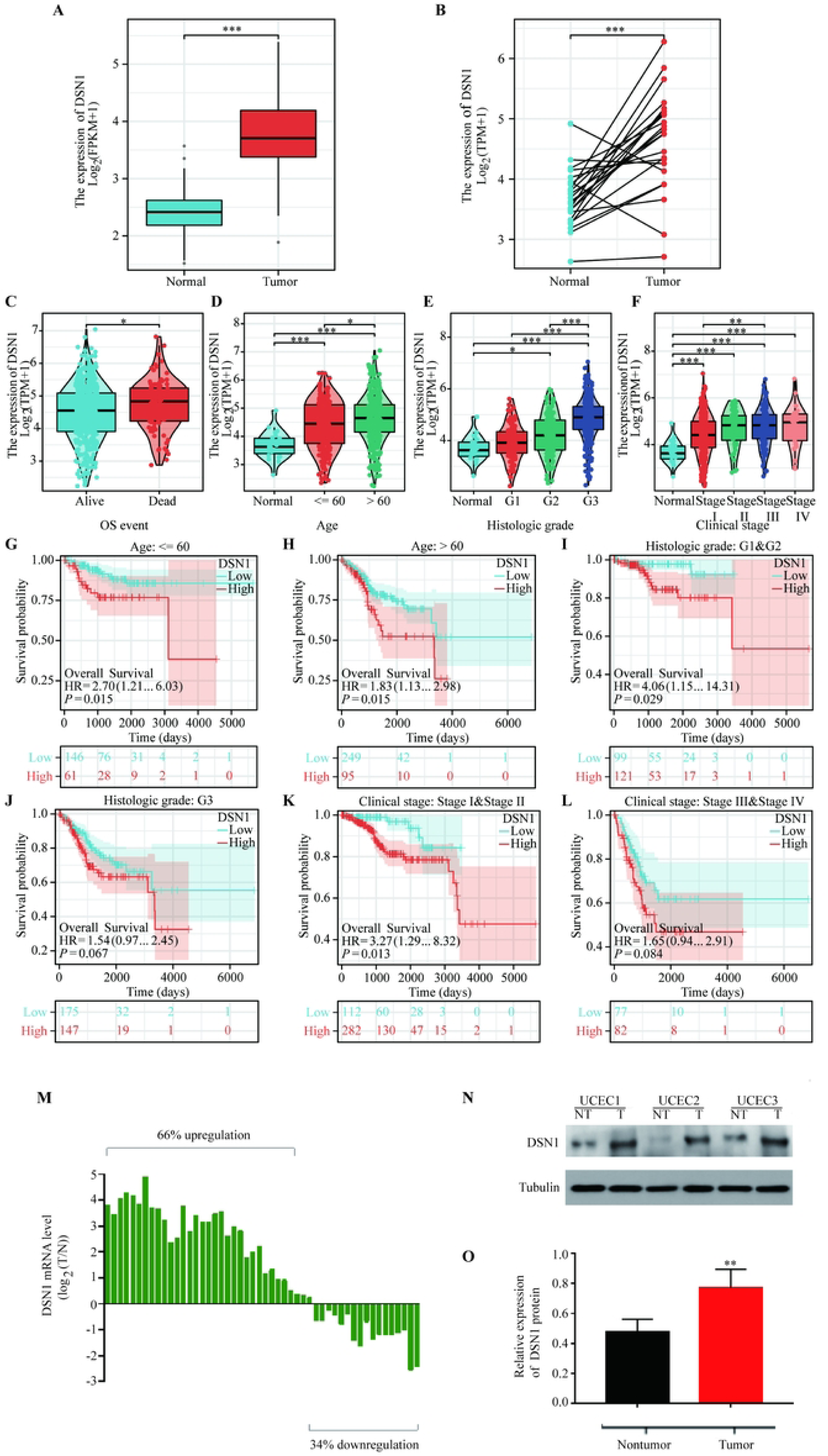
Correlation between DSN1 expression and clinicopathological features. (A) DSN1 mRNA levels in tumor and normal tissues based on the TCGA database. (B) Based on the TCGA database, DSN1 was differentially expressed in UCEC-paired samples. The expression level of DSN1 under the different clinical pathological features, including (C) OS event, (D) age, (E) histologic grade, and (F) clinical stage (*p < 0.05, **p < 0.01, ***p < 0.001). The overall survival curves of patients with differential DSN1 expression under different clinicopathological characteristics include (G) age≤60, (H) ageߣ60, (I) histological grade: G1 and G2, (J) histological grade: G3, (K) clinical stage: stageⅠ and stageⅡ, and (L) stage Ⅲ and stage Ⅳ. (M) Quantitative Real-time PCR analysis of DSN1 mRNA level in 50 cases of UCEC tissues and corresponding normal tissues. Left, a log 2 (T/N) value >0 indicated that DSN1 expression was overexpressed in the UCEC samples; right, a log 2 (T/N) value <0 indicated that DSN1 expression was downregulated in the UCEC samples. (N,O) Determination and quantification of DSN1 protein levels in UCEC tissues and paired non-tumor tissues by western blot.

### Exploring the potential biological functions of DSN1 in UCEC

To study the biological significance of DSNI in UCEC, we found DSN1 co-expression genes (Figure 6A). The hot charts, respectively, showed the top 50 genes that were positively and negatively related to DSN1 (Figure 6B, C). Then, we explored the potential pathways for DSN1 in UCEC. GO analysis suggested that when DSN1 expression was elevated, pathways such as DNA replication were activated (Figure 6D). KEGG analysis showed that when DSN1 expression was elevated, pathways such as the cell cycle, DNA replication, and pyrimidine metabolism were activated (Figure 6E). Meanwhile, GSEA analysis indicated that the elevation of DSN1 expression was closely related to pathways such as the cell cycle, DNA replication, and purine metabolism (Figure 7A-I), showing that DSN1 can affect cell cycle-related pathways.

**Figure 6.**
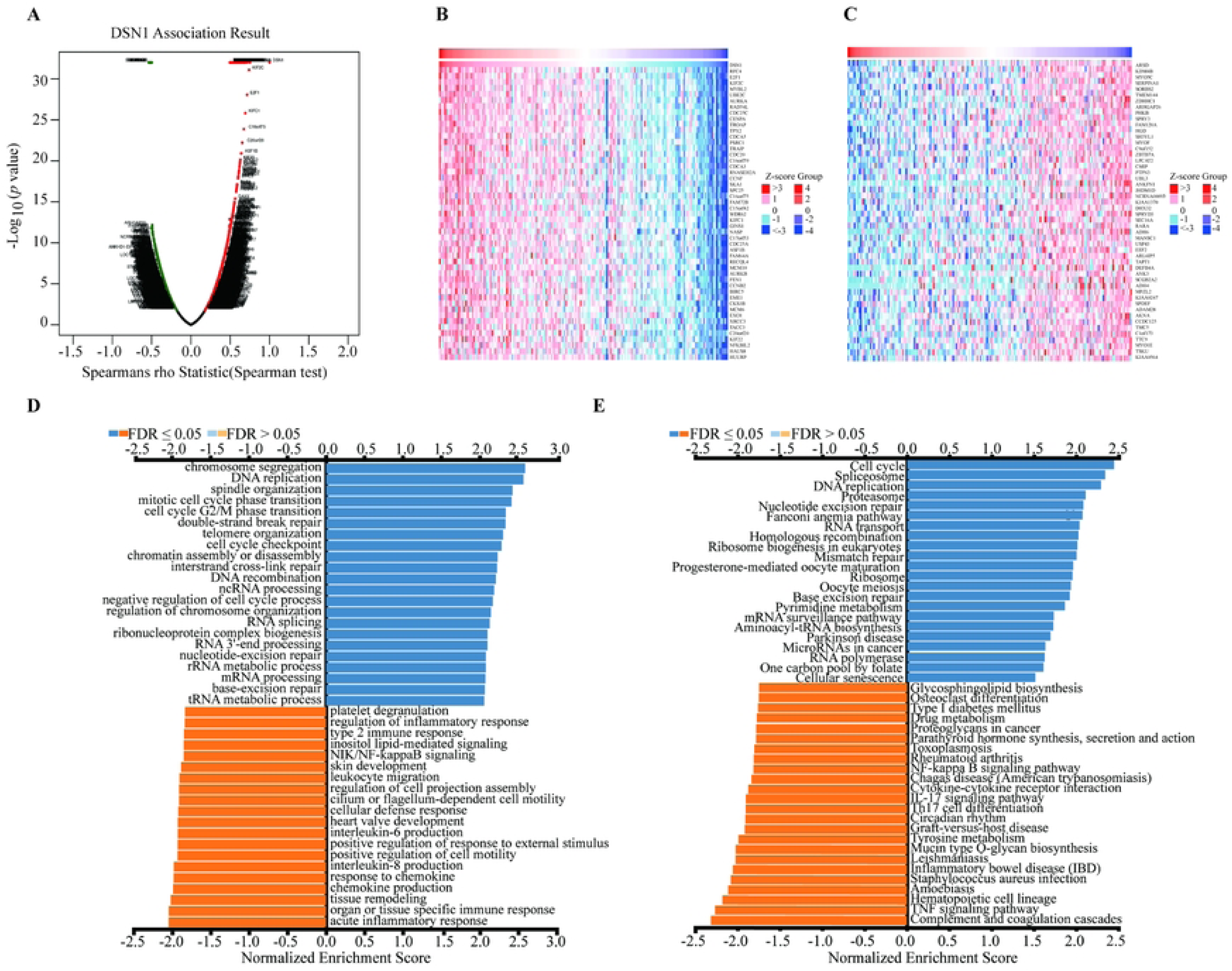
Using Linkedomics to investigate potential biological processes in which DSN1 was involved. (A) Volcano plot of DSN1 co-expressed genes. (B, C) UCEC DSN1 top 50 positive correlation and negative correlation gene heat maps. (D, E) GO and KEGG pathway enrichment analysis of DSN1 in endometrial carcinoma.

**Figure 7.**
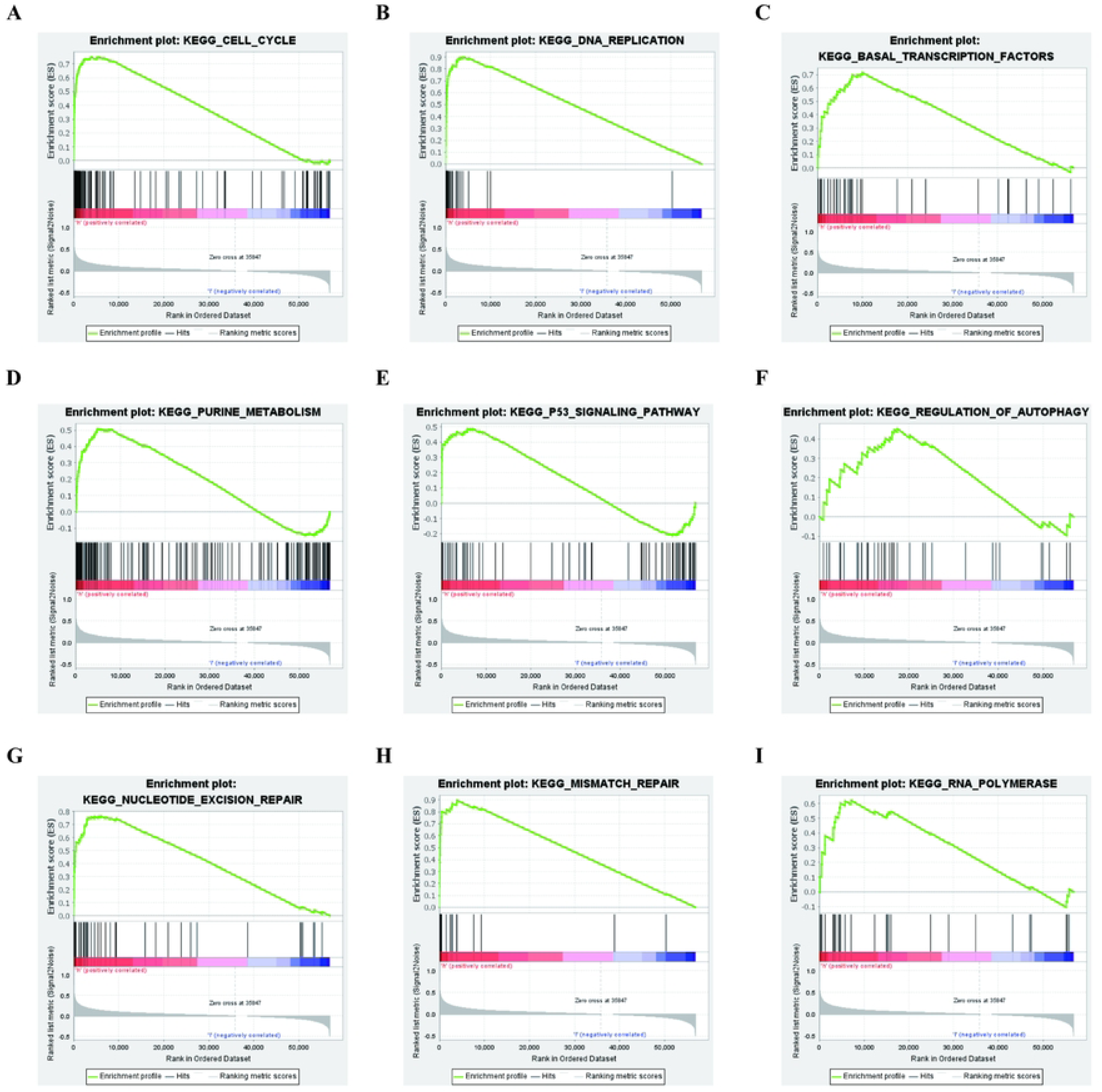
GSEA pathway enrichment analysis. Using GSEA, we found that DSN1 participated in potential biological processes, including (A) cell cycle, (B) DNA replication, (C) basal transcription factors, (D) purine metabolism, (E) P53 signaling pathway, (F) regulation of autophagy, (G) nucleotide excision repair, (H) mismatch repair, and (I) RNA polymerase.

### Construction of PPI networks associated with DSN1

Protein interactions play an irreplaceable role in life processes such as cell cycle regulation. To study the specific mechanisms of DSN1 in cell cycles, we built a network of DNS1 protein interactions (Figure 8A). The first 50 genes of the interaction were shown in Figure 8B. We then screened the top 10 genes of the PPI score for a correlation and survival analysis, showing that all 10 were significantly positive, but only 6 of them had statistically significant survival (Figure 8C, D (Supplementary Figure 2A, B)). Among them, NDC80 has the highest correlation with DSN1, and NDC80 is highly expressed in a variety of cancers, such as liver cancer (13) and epithelial ovarian cancer (14). In addition, NDC80 can also promote the progression of glioma by affecting the cell cycle (15). Therefore, we speculate that DSN1 can influence the UCEC progression by influencing NDC80. WB revealed that NDC80 and Cyclin D1 protein levels dropped when DSN1 was knocked down (Figure 8E). As a result, we postulated that DSN1 may contribute to cancer progression by regulating NDC80 to stimulate the cell cycle.

**Figure 8.**
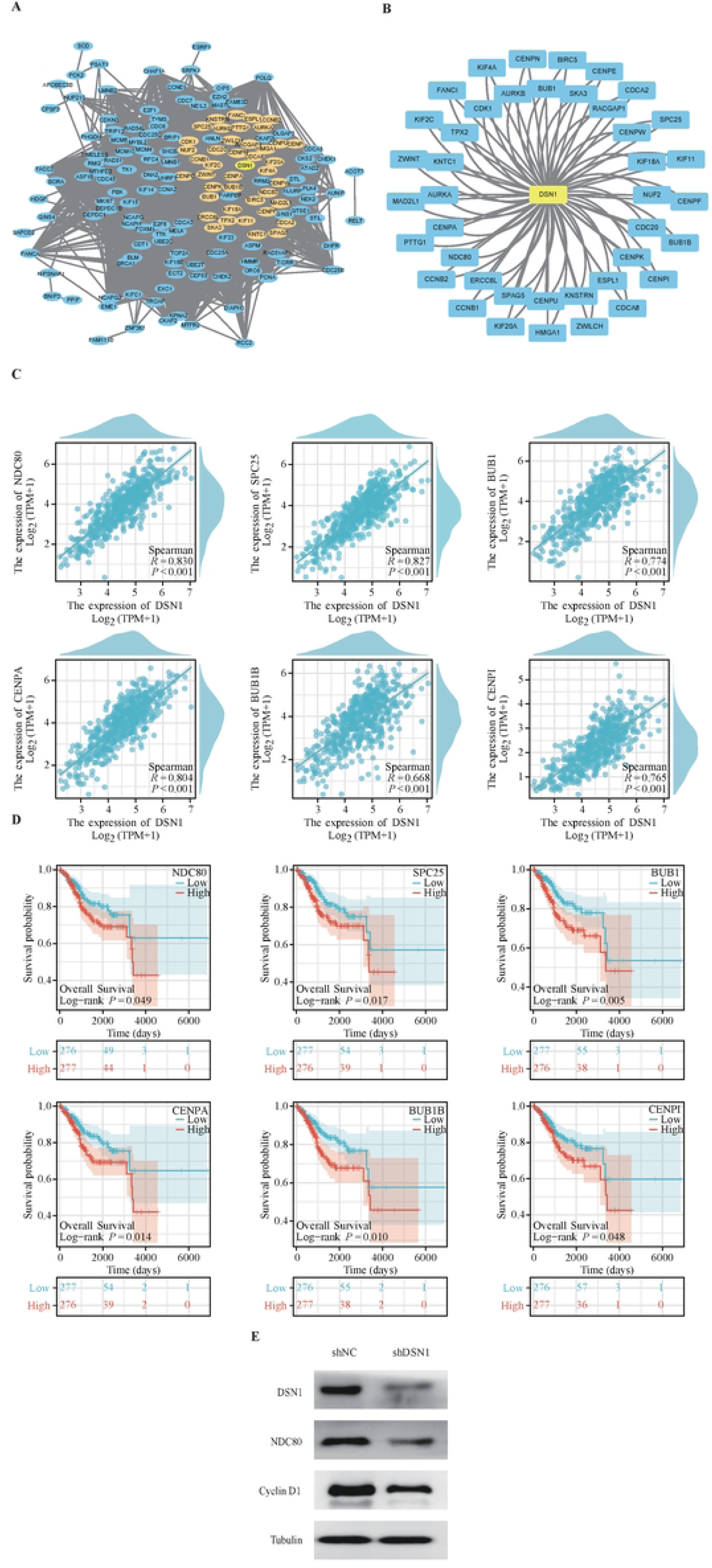
The establishment of the DSN1-related protein-protein interaction network and the analysis of the hub genes. (A, B) DSN1-associated protein-protein interaction (PPI) network. (C) Correlation between DSN1 and the expression of NDC80, SPC25, BUB1, CENPA, BUB1B, and CENPI in UCEC. (D) Prognostic analysis of related genes. (E) Western blot was used to detect DSN1, NDC80, and Cyclin D1 protein expression in UCEC cells stably transfected with the control shRNA or the DSN1 shRNA.

### Relevance of DSN1 expression to immune cell infiltration

To reveal the relationship between DSN1 expression and the UCEC immune response, the immune microenvironment was analyzed between the DSN1 differential expression groups. Multiple immune cells in the DSN1 low-expression group were significantly enriched. For the DSN1 high-expression group, the enrichment of T helper cells, Tgd cells, and Th2 cells was higher (Figure 9A). Simultaneously, it demonstrated that DSN1 was negatively related to the vast majority of immune cells, while it was remarkably positive for Th2 cells, T helper cells, TGD, and Tcm (Figure 9B). Next, we delved into the relationship between DSN1 expression and immune infiltration. It revealed that DSN1 expression was negatively associated with the immune cell infiltration of B cells, CD4+ T cells, and dendritic cells. However, there was a positive correlation with neutrophils and no statistical significance for CD8+T cells and macrophages (Figure 9C). Then, it was found that DSN1 expression was negatively correlated with most chemokines and significantly positively correlated with CCL7 and CCL8 expression in UCEC (Figure 9D, E, F). Furthermore, the Th2 cells and chemokines CCL7 and CCL8 were significantly positively related (Figure 9G, H). Overall, DSN1 may affect the enrichment of Th2 cells in UCEC.

**Figure 9.**
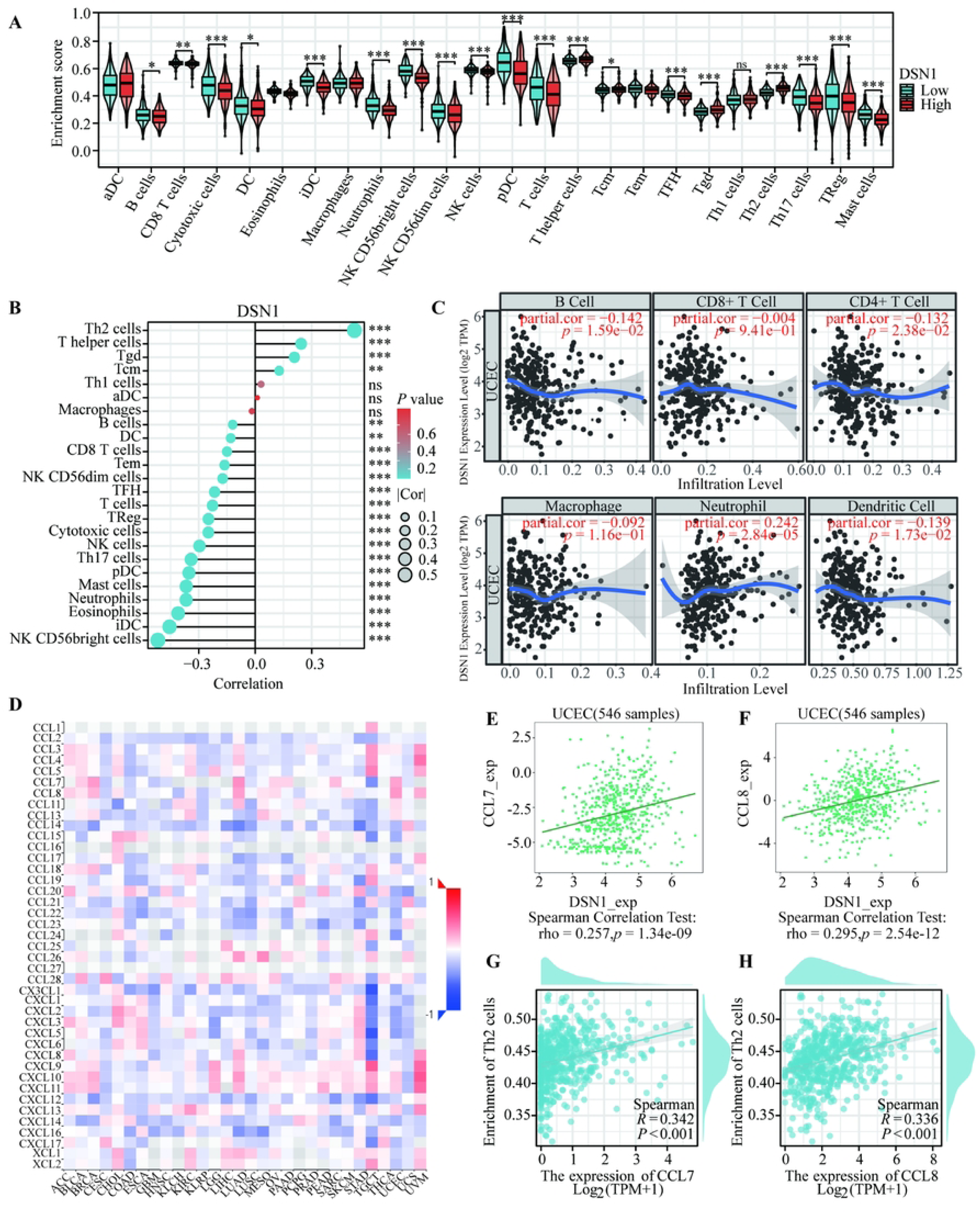
Analysis of the correlation between the DSN1 expression and the immune level and the chemokines. (A) Box plots showed the differences in immune microenvironment between DSN1 high and low expression groups. (B) The lollipops of the correlation of DSN1 expression and immune cells. (C) Correlation of DSN1 expression in UCEC with infiltration levels of B cell, CD8+ T cell, CD4+ T cell, macrophage, neutrophil, and dendritic cell. (D) Heat map of the correlation between DSN1 expression and chemokines in pan-cancer. (E, F) Scatter plot of the correlation between CCL7, CCL8, and DSN1 expression. (G, H) Scatter plots of the correlation between CCL7 and CCL8 expression and Th2 cell enrichment.

### DSN1 drug interaction and drug sensitivity analysis

To explore interactions of DSN1 and cancer treatment drugs, we established drug interaction networks. It showed that seven kinds of drugs could promote the expression of DSN1. However, 11 drugs could inhibit the expression of DSN1 (Figure 10A). We compared cgp2014 and CTD, and cisplatin and sunitinib were found in both them (Figure 10B). IC50 results showed that, when DSN1 had high expression, the cisplatin IC50 was lower, and the sunitinib IC50 was higher than that when DSN1 had low expression (Figure 10C-D). Moreover, it indicated that when DSN1 was highly expressed, the patients were sensitive to cisplatin and were resistant to sunitinib.

**Figure 10.**
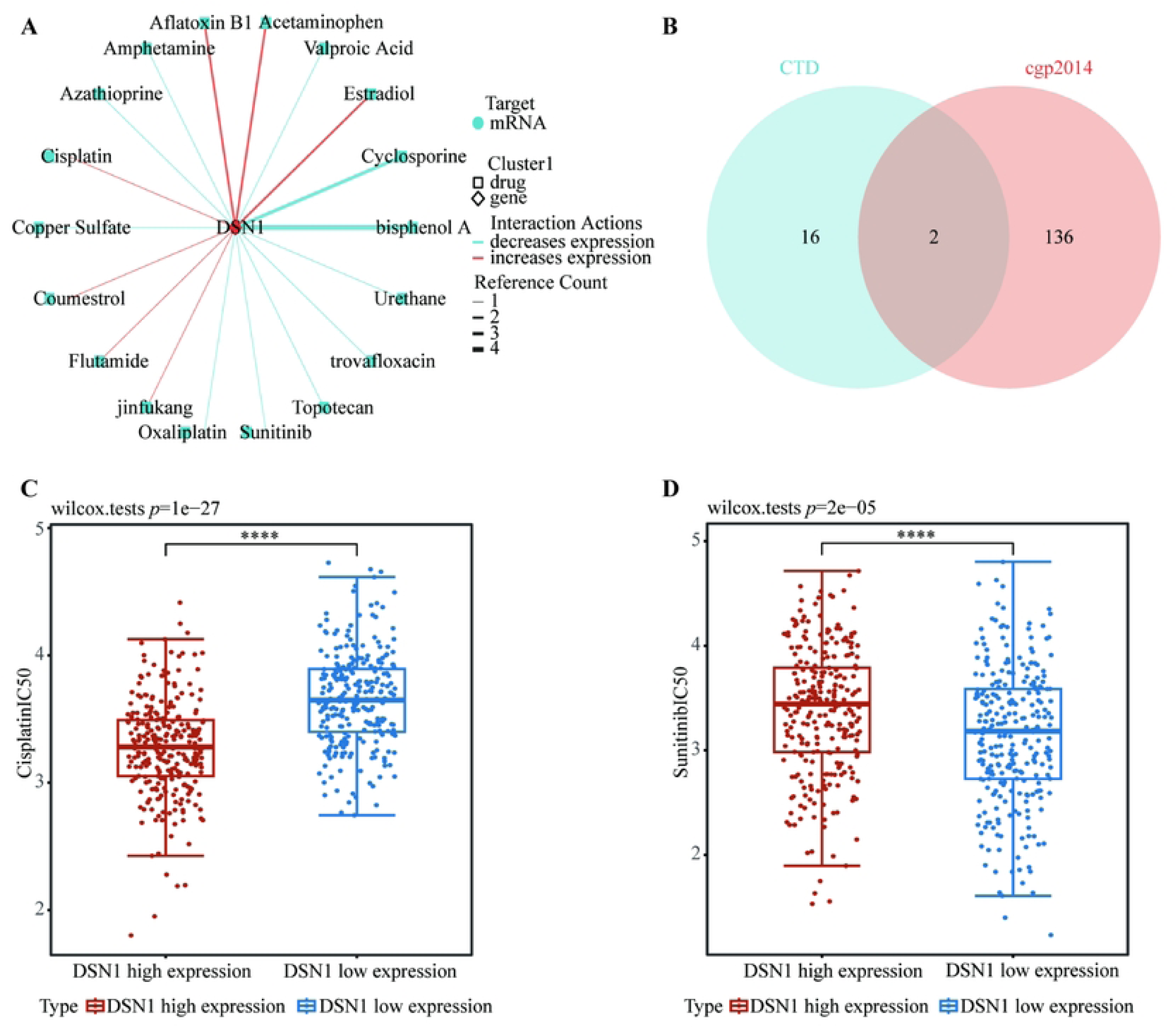
Gene-drug interactions and drug-susceptibility analyses. (A) CTD showed the interaction network between DSN1 and the drug. (B) The Venn figure showed the drug sensitivity results from the two databases. (C-D) The box plots showed the cisplatin IC50 and sunitinib IC50 of DSN1 differential expression.

## Discussion

Endometrial cancer is one of the most prevalent gynecologic malignancies in developed nations. The incidence rate of UCEC in China was 10.54/100,000, and the fatality rate was 2.53/100,000, based on data from the National Cancer Center for the year 2022 (16). In addition, due to the increasing prevalence of obesity and metabolic syndrome, the incidence and mortality of UCEC are still increasing significantly (17). While most patients with early-stage UCEC can be cured with hysterectomy and adjuvant radiotherapy, the prognosis remains poor for patients with late-stage UCEC, with a five-year survival rate of roughly 48% for stage III and 15% for stage IV (18). However, there are currently no biomarkers for the routine clinical diagnosis and prognosis of UCEC (19), so finding new, reliable biomarkers for UCEC and learning more about its molecular biology are urgently needed. In this study, we created a diagnostic model for UCEC by combining multiple databases for comprehensive bioinformatics analysis and experimental verification. We also revealed the expression level and prognostic significance of the key gene DSN1 in the model in UCEC and further explored the biological mechanism of DSN1 promoting the occurrence and progression of UCEC.

In this study, we combined three endometrial cancer mRNA microarray datasets from the GEO database (GSE17025, GSE39099, and GSE106191) and the endometrial cancer datasets from the TCGA database for differential gene analysis. We then performed WGCNA analysis of these differential genes and obtained 164 core genes that were significantly associated with UCEC in both databases. Then, through pathway enrichment analysis, we found that these 164 core genes were substantially enriched in cell cycle-related pathways, such as mitotic cell cycle transition, regulation of cell cycle transition, DNA replication, and other signaling pathways. At the same time, through random forest analysis, the top 15 genes with the highest score among 164 core genes were found. We then performed lasso regression analysis on these 164 genes and obtained 20 core genes for UCEC diagnosis. In addition, we also used consensus cluster analysis to divide UCEC into three major subtypes based on these 20 genes and constructed associated prognostic models.

Then, through the survival analysis of these 20 genes and taking into account the random forest algorithm mentioned above, DSN1 can influence patient prognosis in addition to having a major impact on the random forest algorithm. At the same time, DSN1, as a kinetochore protein-coding gene, is widely distributed in centromere (20) and is crucial for preserving the stability of centromere structure and proper chromosome separation (10). Previous studies have shown that DSN1 can alter the cell cycle, which can accelerate the growth of certain malignancies. For example, in colorectal cancer, DSN1 can be stabilized in an M6A-related manner that promotes cancer progression (21); in breast cancer, DSN1 expression is significantly upregulated and is strongly linked to a poor prognosis and decreased survival (7). However, the expression of DSN1 in endometrial cancer and its biological function remain unclear. Our investigation revealed a correlation between the prognosis and expression level of DSN1 and several clinicopathological characteristics of UCEC, such as age, histological grade, and clinical stage. Further, we also confirmed that the expression of DSN1 in endometrial carcinoma tissues was greater than that in normal control tissues by qRT-PCR and western blot, which was consistent with bioinformatics analysis. In summary, DSN1’s mRNA and protein levels in UCEC have changed, but more research is still needed to determine the precise mechanism of action.

In order to further reveal the specific biological mechanism of DSN1 affecting UCEC, we conducted the enrichment analysis of the GOKEGG pathway and the GSEA pathway. The outcomes demonstrated that DSN1 was considerably activated in cell cycle, DNA replication, chromosome separation, and other pathways. It is well known that cell cycle is one of the main cellular mechanisms influencing the development of malignant tumors (22) and having a significant impact on metastasis, immunity, and tumor metabolism (23). The high expression of DSN1 in UCEC may promote the growth of tumor cells by influencing the cell cycle. We further verified this hypothesis through experiments. The experimental results demonstrated that Cyclin D1 protein level decreased following DSN1 knockdown. These results suggest that DSN1 knockdown can inhibit the cell cycle, thereby inhibiting tumor growth. In summary, our study revealed that DSN1 is likely to promote cell proliferation and cancer progression in UCEC by promoting the UCEC cell cycle.

Subsequently, through PPI analysis, we found that DSN1 had protein interaction with NDC80, SPC25, BUB1, CENPA, BUB1B, and CENPI. Among them, NDC80 is the gene with the highest correlation with DSN1, and as the outer component of the centromere and the regulator of spindle check points, NDC80 is highly expressed in a variety of cancers. NDC80 knockdown in pancreatic cancer can cause cell cycle halt, which slows tumor development (24). NDC80 knockdown in gastric cancer can significantly reduce cell growth in vitro and in vivo, preventing cell division during the G2/M phase (25). Furthermore, research has shown that NDC80 is significantly overexpressed in endometrial cancer and is associated with a poor prognosis (26). Thus, we hypothesize that DSN1’s interaction with NDC80 may have an impact on the onset and progression of UCEC. Through experiments, we found that in the shDSN1 group, NDC80 and Cyclin D1 protein levels were significantly reduced. Through these experiments, we have reason to believe that DSN1 affects the cell cycle through NDC80 and thus mediates the incidence and growth of tumors.

Given the growing body of research demonstrating the pivotal role the tumor microenvironment plays in tumor initiation, development, metastasis, and response to therapy (27), we looked at the relationship between DSN1 expression and immune cell infiltration in UCEC. The results demonstrated a negative correlation between DSN1 expression and the majority of immune cells’ infiltration abundance and a positive correlation with Th2 cells’ infiltration abundance. Th2 cells have the function of promoting tumor progression and immunosuppression, which is often linked to a poor prognosis of various tumors (28), and Th1/Th2 balance restoration is crucial in the therapy of cancers (29). Then, we examined the chemokines linked to DSN1 expression to look into the connection between DSN1 and immune cell infiltration in more detail. The findings demonstrated that DSN1 was negatively correlated with most chemokines in UCEC but positively correlated with CCL7 and CCL8. And CCL7 and CCL8 were also positively correlated with Th2 cells in UCEC. According to previous studies, CCL7 can enrich Th2 cells during scar formation, thus affecting the local expression levels of Th1 and Th2 cells (30). In vitiligo, CCL8 can deflect T cells toward pathogenic Th2 cells (31). Therefore, we speculate that DSN1 is likely to affect the infiltration level of Th2 cells by regulating CCL7 and CCL8, thus promoting tumor progression and immunosuppression. In summary, taking advantage of the properties of DSN1 that may increase the proportion of Th2 cells, it is possible to try to combine DSN1 inhibitors with immune checkpoint inhibitors to further enhance the effect of immunotherapy in UCEC patients.

In conclusion, our study integrated various bioinformatics approaches across multiple datasets to ultimately establish a diagnostic and prognostic model for UCEC, in which DSN1 is expected to be a biomarker for early diagnosis and treatment of endometrial cancer. Bioinformatics analysis and experiments confirmed that DSN1 was overexpressed in UCEC and correlated with some clinicopathological features. At the same time, we also tested how DSN1 affects the cell cycle through experiments. Finally, we proposed and predicted the interaction of DSN1 with NDC80, as well as analyzed drugs that were sensitive to cells with high DSN1 expression.

However, we need to acknowledge several limitations related to this study. First, we obtained data on UCEC mainly from online public databases, and larger studies are still needed in the future to validate and strengthen our findings. Secondly, we only verified the interaction of DSN1 with NDC80 and did not further verify their specific mode of action through experiments.

## Data Availability

All datasets in the present study were downloaded from public databases. These public databases allowed researchers to download and analyze public datasets for scientific purposes.

## Acknowledgements

We thank the mentioned public databases for providing us the data and analytical tools.

## Supplementary Figure captions

**Supplementary Figure 1. The survival curves of 12 genes for which they were not statistically significant.** (A) 12 genes with no statistically significant survival curves.

**Supplementary Figure 2. Correlation analysis and survival analysis of four genes for which survival analysis was not meaningful**. The correlation scatter plot for genes without survival statistical significance for (A) AURKB, MAD2L1, NUF2, and ZWINT. The survival curves of the genes include (B) AURKB, MAD2L1, NUF2, and ZWINT.

